# Gene-level germline contributions to clinical risk of recurrence scores in Black and White breast cancer patients

**DOI:** 10.1101/2021.03.19.21253983

**Authors:** Achal Patel, Montserrat García-Closas, Andrew F. Olshan, Charles M. Perou, Melissa A. Troester, Michael I. Love, Arjun Bhattacharya

## Abstract

Continuous risk of recurrence scores (CRS) based on tumor gene expression are vital prognostic tools for breast cancer (BC). Studies have shown that Black women (BW) have higher CRS than White women (WW). Although systemic injustices contribute substantially to BC disparities, evidence for biological and germline contributions is emerging. We investigated germline genetic associations with CRS and CRS disparity using approaches modeled after transcriptome-wide association studies (TWAS). In the Carolina Breast Cancer Study, using race-specific predictive models of tumor expression from germline genetics, we performed race-stratified (N=1,043 WW, 1083 BW) linear regressions of three CRS (ROR-S: PAM50 subtype score; Proliferation Score; ROR-P: ROR-S plus Proliferation Score) on imputed Genetically-Regulated tumor eXpression (GReX). Using Bayesian multivariate regression and adaptive shrinkage, we tested GReX-prioritized genes for associations with PAM50 tumor expression and subtype to elucidate patterns of germline regulation underlying GReX-CRS associations. At FDR-adjusted *P* < 0.10, we detected 7 and 1 GReX-prioritized genes among WW and BW. Among WW, CRS were positively associated with *MCM10, FAM64A, CCNB2*, and *MMP1* GReX and negatively associated with *VAV3, PCSK6*, and *GNG11* GReX. Among BW, higher *MMP1* GReX predicted lower Proliferation score and ROR-P. GReX-prioritized gene and PAM50 tumor expression associations highlighted potential mechanisms for GReX-prioritized gene to CRS associations. Among BC patients, we find differential germline associations with CRS by race, underscoring the need for larger, diverse datasets in molecular studies of BC. Our findings also suggest possible germline *trans*-regulation of PAM50 tumor expression, with potential implications for CRS interpretation in clinical settings.

**SIGNIFICANCE:** We find race-specific genetic associations with breast cancer risk-of-recurrence scores (CRS). Follow-up analyses suggest mediation of these associations by PAM50 molecular subtype and gene expression, with implications for clinical interpretation of CRS.

## INTRODUCTION

Tumor expression-based molecular profiling has improved clinical classification of breast cancer (1-3). One tool is the PAM50 assay, which integrates tumor expression of 50 genes (derived from a set of 1,900 subtype-specific genes identified in microarray studies) to determine PAM50 intrinsic molecular subtypes: Luminal A (LumA), Luminal B (LumB), Human epidermal growth factor 2-enriched (HER2-enriched), Basal-like, and Normal-like (1,4). Intrinsic molecular subtypes are strong prognostic factors for breast cancer outcomes, including recurrence and mortality. For instance, Basal-like breast cancer has substantially higher recurrence and mortality risk compared to LumA breast cancer (5-8). In recent years, continuous risk of recurrence scores (CRS) have gained traction as a potential clinical tool that encapsulates prognostic differences of breast cancer intrinsic molecular subtypes into a singular measure that can be used to guide treatment decisions. CRS include ROR-S, Proliferation score, ROR-P, and ROR-PT (1,9). ROR-P, for instance, is determined by combining ROR-S (PAM50 tumor expression-based subtype score) and Proliferation score (tumor expression of 11 PAM50 genes). ROR-PT further integrates ROR-P with information on tumor size. Studies show that CRS offer significant prognostic information beyond clinical variables (e.g., nodal status, tumor grade, age, hormonal therapy), improve adjuvant treatment decisions, and offer robust risk stratification for distant (5-10 years post diagnosis) recurrence (10-12).

In the Carolina Breast Cancer Study (CBCS), Black women (BW) with breast cancer have disproportionately higher CRS than White Women (9), and similar disparities have been noted in Oncotype Dx recurrence score (9,13). Systemic injustices, like disparities in healthcare access, explain a substantial proportion of breast cancer outcome disparities (14-17). Recent studies additionally suggest that germline genetic variation is associated with breast cancer outcomes, and these associations vary across ancestry groups (18-21). In The Cancer Genome Atlas (TCGA), BW had substantially higher polygenic risk scores for the more aggressive ER-negative subtype than WW, suggesting differential genetic contributions for susceptibility for breast cancer, especially ER-negative breast cancer (21). In a transcriptome-wide association study (TWAS) of breast cancer mortality, germline-regulated gene expression (GReX) of four genes was associated with mortality among BW and gene expression for no genes was associated among WW (18). However, the role of germline genetic variation in recurrence, CRS, and CRS disparity remains a critical knowledge gap. Studying genetic associations with breast cancer outcomes in BW is necessary to ensure advancements in breast cancer genetics are not limited to or generalizable in only White populations, thus aiding in decreasing health disparities.

As racially-diverse genetic datasets typically have small samples of BW, gene-level association tests can increase study power. These approaches include TWAS, which integrates relationships between single nucleotide polymorphisms (SNP) and gene expression with genome-wide association studies (GWAS) to prioritize gene-trait associations (22,23). TWAS aids in interpreting genetic associations by mapping significant GWAS associations to tissue-specific expression of individual genes. In cancer applications, TWAS has identified susceptibility genes at loci previously undetected through GWAS, highlighting its improved power and interpretability (24-26). Previous studies show that stratification of the entire TWAS (model training, imputation, and association testing) is preferable in diverse populations, as models may perform poorly across ancestry groups and methods for TWAS in admixed populations are unavailable (18,27).

Here, using data from the CBCS, which includes a large sample of Black breast cancer patients with tumor gene expression data, we study race-specific germline genetic associations for CRS using a gene-based association testing approach that borrows from TWAS methodology. CRS included in this study are ROR-S, Proliferation score, and ROR-P. Using race-specific predictive models for tumor expression from germline genetics, we identify sets of GReX-prioritized genes (i.e. genes whose GReX is associated with CRS) across BW and WW. We additionally investigate ROR-P specific GReX-prioritized genes for associations with PAM50 subtype and subtype-specific tumor gene expression to elucidate germline contributions to PAM50 subtype, and how these mediate GReX-prioritized gene and CRS associations. Unlike previous studies that correlated tumor gene expression (as opposed to germline-regulated tumor gene expression) with subtype or subtype-specific tumor gene expression, TWAS enables directional interpretation of observed associations (22,23).

## MATERIALS AND METHODS

### Data collection

#### Study population

The CBCS is a population-based study of North Carolina (NC) breast cancer patients, enrolled in three phases; study details have been previously described (28,29). Patients aged 20 to 74 were identified using rapid case ascertainment with the NC Central Cancer Registry with randomized recruitment to oversample self-identified Black and young women (ages 20-49) (9,29). Demographic and clinical data (age, menopausal status, body mass index, hormone receptor status, tumor stage, study phase, recurrence) were obtained through questionnaires and medical records. The study was approved by the Office of Human Research Ethics at the University of North Carolina at Chapel Hill, and informed consent was obtained from each participant.

#### CBCS genotype data

Genotypes were assayed on the OncoArray Consortium’s custom SNP array (Illumina Infinium OncoArray) (30) and imputed using the 1000 Genomes Project (v3) as a reference panel for two-step phasing and imputation using SHAPEIT2 and IMPUTEv2 (31-34). The DCEG Cancer Genomics Research Laboratory conducted genotype calling, quality control, and imputation (30). We excluded variants with less than 1% minor allele frequency and deviations from Hardy-Weinberg equilibrium at *P* < 10^−8^ (35,36). We intersected genotyping panels for BW and WW samples, resulting in 5,989,134 autosomal variants and 334,391 variants on the X chromosome (37). We only consider the autosomal variants in this study.

#### CBCS gene expression data

Paraffin-embedded tumor blocks were assayed for gene expression of 406 breast cancer-related and 11 housekeeping genes using NanoString nCounter at the Translational Genomics Laboratory at UNC-Chapel Hill (4,9). These 406 breast cancer-related genes include genes part of the PAM50, P53, E2, IGF, and EGFR signatures, among others (**Supplementary Table S1**). As described previously, we eliminated samples with insufficient data quality using NanoStringQCPro (18,38), scaled distributional difference between lanes with upper-quartile normalization (39), and removed two dimensions of unwanted technical and biological variation, estimated from housekeeping genes using RUVSeq (39,40). The current analysis included 1,199 samples with both genotype and gene expression data (628 BW, 571 WW).

### Statistical analysis

#### Overview of GReX and TWAS

We adopted TWAS methodology to construct GReX (exposure of interest in this study). GReX for a given gene represents the portion of tumor expression explained by *cis*-genetic regulation; GReX was constructed for the aforementioned set of BC-related genes (**Supplementary Table S1**). Briefly, TWAS integrates expression data with GWAS to prioritize gene-level germline-trait associations through a two-step analysis (**Figure 1A-BW**). First, using germline and transcriptomic data, we trained predictive models of tumor gene expression using all SNPs within 0.5 Megabase of the gene (18,23). Second, we used these models to impute the GReX of a gene by multiplying the SNP-gene weights from the predictive model with the dosages of each SNP. Associations between GReX (for a given gene) and trait (CRS, for instance) in regression analyses identify gene-trait relationships that are a consequence of germline variation. If sufficiently heritable genes are assayed in the correct tissue, TWAS-based GReX analyses increase power to detect germline-trait associations and aids interpretability of results, as associations are mapped from germline genetics to individual genes (23,41).

**Figure 1.**
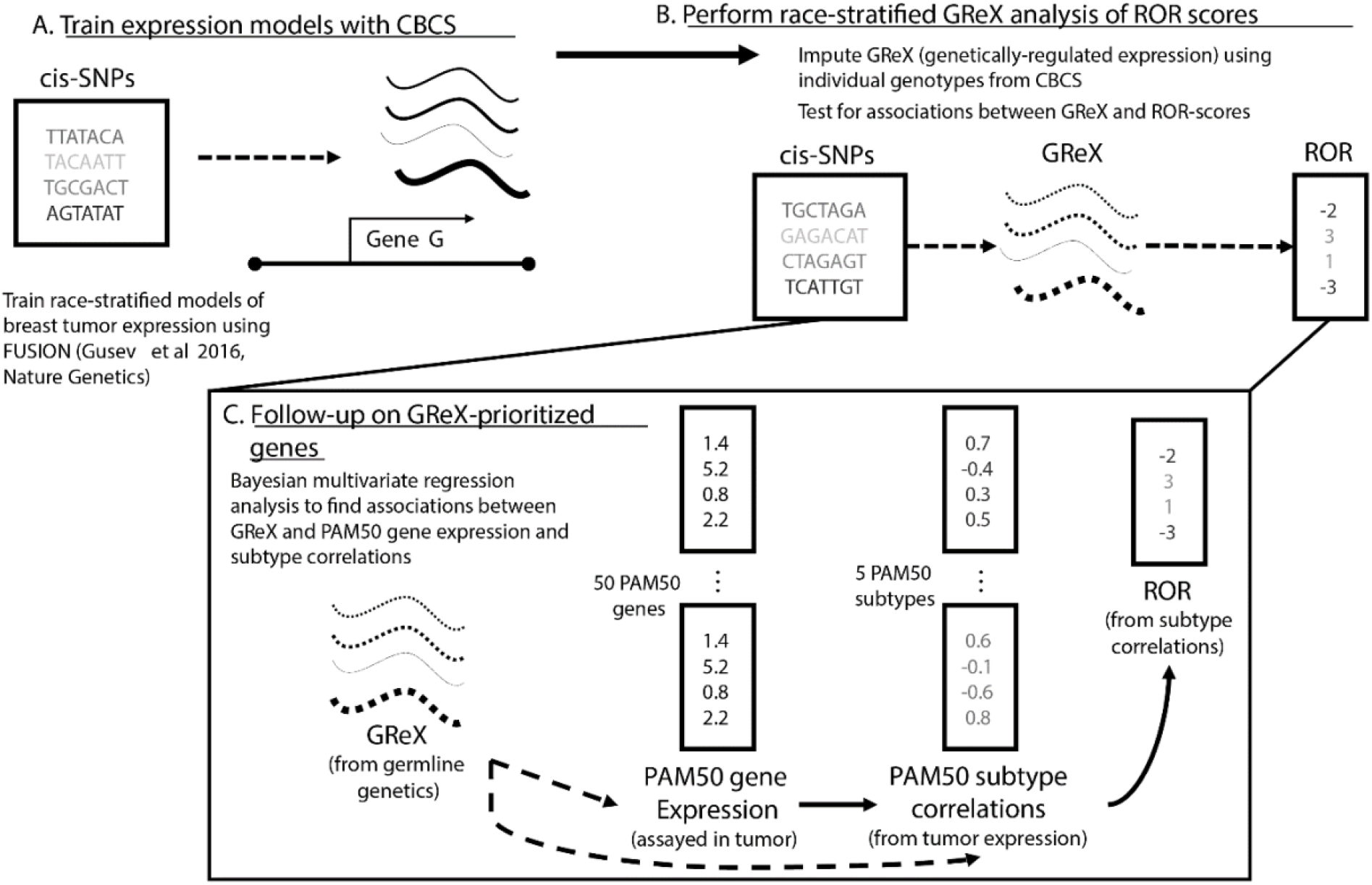
Schematic of study analytic approach. A) In CBCS, constructed race-stratified predictive models of tumor gene expression from *cis*-SNPs. B) In CBCS, imputed GReX at individual-level using genotypes and tested for associations between GReX and CRS in race-stratified linear models; only GReX of genes with significant *cis*- h^2^ and high cross validation performance (*R*^2^ > 0.01 between observed and predicted expression) considered for race-stratified association analyses. C) Follow-up analyses on GReX-prioritized genes (i.e., genes whose GReX were significantly associated with CRS at FDR <0.10). In race-stratified models, PAM50 SCCs and PAM50 tumor expressions were regressed against GReX-prioritized genes under a Bayesian multivariate regression and multivariate adaptive shrinkage approach.

#### GReX analysis of CRS in CBCS

We adopted techniques from FUSION to train predictive models of tumor expression from *cis*-germline genotypes (18,23). Motivated by strong associations between germline genetics and tumor expression in CBCS (18), for genes with non-zero *cis*-heritability at nominal *P* < 0.10, we trained predictive models for covariate-residualized tumor expression with all *cis*-SNPs within 0.5 Megabase using linear mixed modeling or elastic net regression (**Supplementary Methods, Supplementary Materials**) (42,43). Here, we used the 628 BW samples and 571 WW samples with both genotype and expression data to train these race-specific expression models. We selected models with five-fold cross-validation adjusted *R*^2^ > 0.01 between predicted and observed expression values, resulting in 59 and 45 models for WW and BW, respectively. Further details on these models, including heritability and cross-validation performance are available at **Supplementary Table S2**. These models also showed sufficiently strong predictive performance in external validation using TCGA data (18).

Using only germline genetics as input, we imputed GReX in 1,043 WW and 1,083 BW, respectively, in CBCS. For samples not present in the training dataset, we multiplied the SNP weights from the predictive models with the SNP dosages to construct GReX. For samples in both the training and imputation datasets, GReX was imputed via cross-validation to minimize data leakage. We tested GReX for associations with ROR-S, Proliferation Score, and ROR-P using multiple linear regression adjusted for age, estrogen receptor (ER) status, tumor stage, and study phase (1). We corrected for test-statistic bias and inflation using a Bayesian bias and inflation adjustment method *bacon*, as TWAS are prone to bias and inflation of test statistics (44). We then adjusted for multiple testing using the Benjamini-Hochberg procedure (44,45). As a comparison for the germline effect of GReX-prioritized genes, we additionally assessed the effect of total (germline-regulated and post-transcriptional) tumor expression of those GReX-prioritized genes on CRS using similar linear models. We were underpowered to study time-to-recurrence, as recurrence data was collected only in CBCS Phase 3 (635 WW, 742 BW with GReX and recurrence data; 183 WW, 283 BW with tumor expression and recurrence data). For significant GReX-prioritized genes for CRS (FDR-adjusted P < 0.10), we conducted follow-up permutation tests: we shuffle the SNP-gene weights in the predictive model 5,000 times to generate a null distribution and compare the original GReX-CRS associations to this null distribution. This permutation test assessed whether the GReX association provides more tissue-specific expression context, beyond any strong SNP-CRS associations at the genetic locus (23).

#### PAM50 assay and ROR-S, Proliferation score, and ROR-P calculation

As described previously (1), using partition-around-medoid clustering, we calculated the correlation with each subtype’s centroid for study individuals based on PAM50 expressions (10 PAM50 genes per subtype). The largest subtype-centroid correlation defined the individual’s molecular subtype. ROR-S was determined via a linear combination of the PAM50 subtype-centroid correlations (SCCs); the coefficients to the PAM50 SCCs in the linear combination are positive for Luminal B, HER2-enriched, and Basal-like and negative for Luminal A (1). Proliferation score was computed using log-scale expression of 11 PAM50 genes, while ROR-P was computed by combining ROR-S and Proliferation score.

Assignment of PAM50 gene to subtype was based on PAM50 gene centroid values for each subtype; a PAM50 gene is assigned to the subtype with the largest positive centroid value. Subtype assignment through this “greedy algorithm” are specific to this study and represent a simplified reality (e.g., ESR1 classified as part of Luminal A subtype only even though *ESR1* expression correlates with both Luminal A and to a slightly lesser degree Luminal B subtype). Moreover, subtype assignment for this portion of analyses was conducted only for visual comparison of patterns of associations between GReX-prioritized genes and PAM50 tumor gene expressions (i.e., subtype assignment in this portion of analyses had no bearing on continuous ROR score calculations or subtype-centroid correlations).

#### Bayesian multivariate regressions and multivariate adaptive shrinkage

As previously noted (1), CRS are functions of PAM50 SCCs and gene expression profiles. Thus, we followed up on CRS-associated GReX-prioritized genes by studying their associations with PAM50 SCCs and gene expression. We assessed GReX-prioritized genes (for ROR-P) in relation to SCCs and PAM50 tumor gene expression (**Figure 1C**). Importantly, consistent with the original formulation of ROR-S, we did not consider normal-like subtype and normal-like subtype specific genes; subtype-specific genes were determined using a greedy assignment algorithm, described in the previous section. This classification scheme offers analytic simplicity but is an oversimplification for some PAM50 genes. We found that none of our GReX-prioritized genes were within 1 Megabase of PAM50 genes and that most GReX-prioritized genes were not on the same chromosome as PAM50 genes (**Supplementary Table S3**).

Existing gene-based mapping techniques for *trans*-expression quantitative trait loci (eQTL) (SNP and gene are separated by more than 1 Megabase) mapping include *trans*-PrediXcan and GBAT (46,47). We employed Bayesian multivariate linear regression (BtQTL) to account for correlation in multivariate outcomes (SCCs and PAM50 gene expression) in association testing. BtQTL improves power to detect significant *trans*-associations, especially when considering multiple genes with highly correlated (>0.5) expression (**Supplementary Figures S1-S2**). Lastly, we conducted adaptive shrinkage on BtQTL estimates using mashr, an empirical Bayes method to estimate patterns of similarity and improve accuracy in associations tests across multiple outcomes (48). mashr outputs revised posterior means, standard deviations, and corresponding measures of significance (local false sign rates, or LFSR).

*Associations of genetic ancestry and race with tumor expression and GReX of GReX-prioritized genes* Prior studies using CBCS have reported concordance between self-reported race and genetic ancestry (first principal component of combined genotype matrix) (49). In an effort to further contextualize CRS associations across race and to disentangle race from genetic ancestry in our study population (specifically, whether race, which captures both genetic ancestry and socioeconomic context, is a proxy for genetic ancestry in our study population), we investigated: 1) association between genetic ancestry and tumor expression of GReX-prioritized genes; 2) association between genetic ancestry and GReX of GReX-prioritized genes; 3) association between race and tumor expression of GReX-prioritized genes; 4) association between race and GReX of GReX-prioritized genes. Genetic ancestry was computed by aggregating across local ancestry, as determined through the RFMix pipeline (50).

## RESULTS

### Race-specific associations between GReX and CRS

We performed race-specific GReX analysis for CRS to investigate the role of germline genetic variation in CRS and CRS racial disparity. We identified 8 genes (*MCM10, FAM64A, CCNB2, MMP1, VAV3, PCSK6, NDC80, MLPH)*, 8 genes (*MCM10, FAM64A, CCNB2, MMP1, VAV3, NDC80, MLPH, EXO1)*, and 10 genes (*MCM10, FAM64A, CCNB2, MMP1, VAV3, PCSK6, GNG11, NDC80, MLPH, EXO1)* whose GReX was associated with ROR-S, proliferation, and ROR-P, respectively, in WW, and 1 gene (*MMP1*) whose GReX was associated with proliferation and ROR-P in BW at FDR-adjusted *P* < 0.10 (**Figure 2A, 2B**). No associations were detected between GReX and ROR-S among BW. We refer to genes with statistically significant GReX analysis associations (FDR-adjusted P < 0.10) as GReX-prioritized genes. Among these identified genes, only genes that are not part of the PAM50 panel (i.e., excluding *NDC80, MLPH, EXO1*) were considered in downstream permutation and GReX-prioritized gene follow up analyses (**Figure 1C**), as we wished to focus investigation on relationship between non-PAM50 GReX-prioritized genes and PAM50 (tumor) genes. **Supplementary Figure S3** shows results from a sensitivity analysis comparing the effect sizes for the GReX-CRS associations within samples used in training, not used in training, and the overall associations using all training and non-training samples. In general, we see concordance in the direction of association across these three splits of data, though some of the associations detected within only training or non-training samples intersect the null.

**Figure 2.**
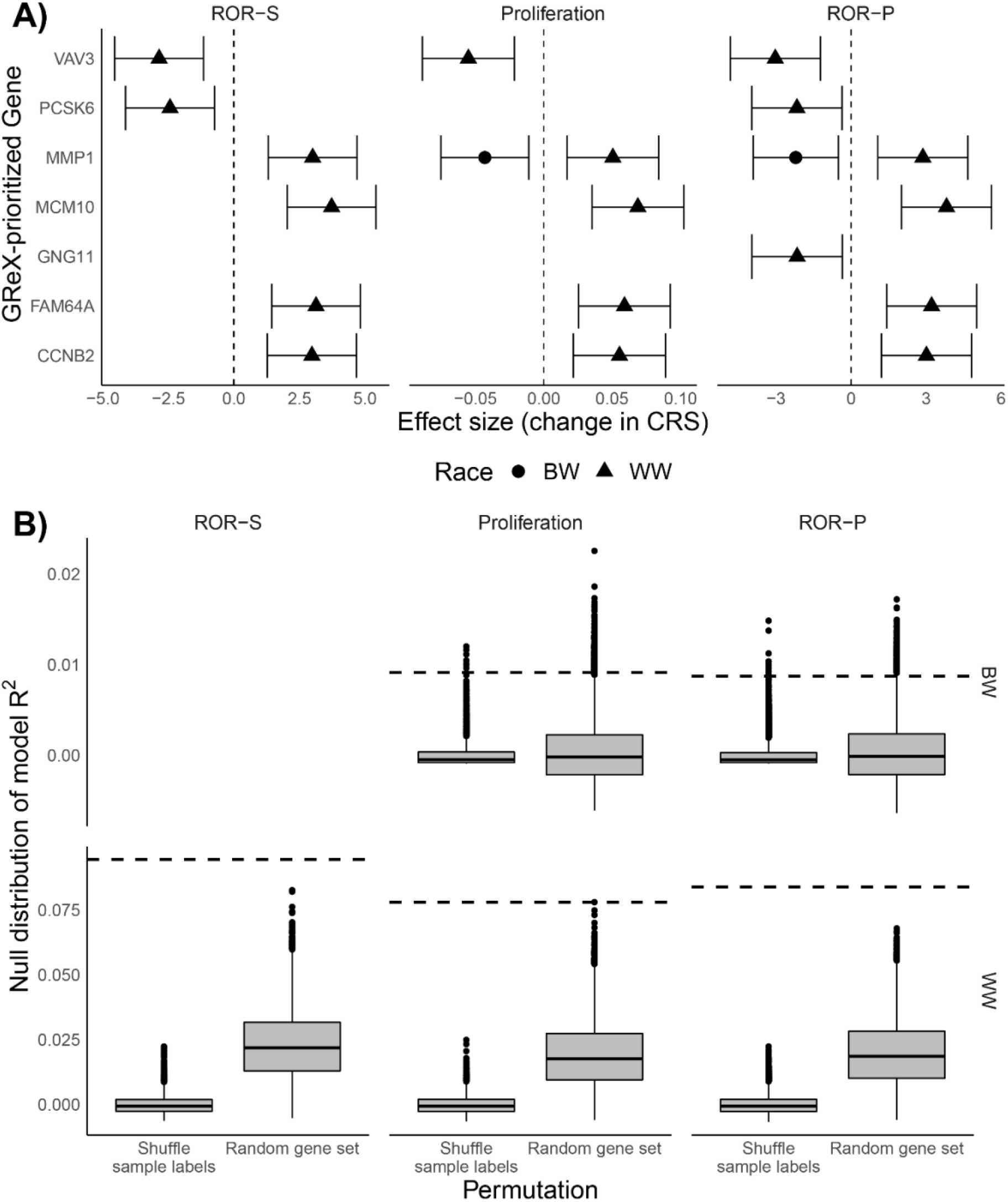
Permutation tests and associations between GReX-prioritized genes and CRS for WW and BW. A) Effect estimates correspond to change in ROR-S, Proliferation score, and ROR-P per one standard deviation increase in GReX-prioritized gene expression (i.e., one standard deviation increase in GReX of gene). Triangle denotes WW and circle denotes BW. B) Boxplots correspond to null distributions (shuffled GReX-sample labels on left, random set of genes on right) of covariates residualized-*R*^2^ for regressions of CRS on GReX-prioritized genes. Null distributions are provided for 10,000 permutations of the GReX-sample labels and 10,000 random sets of genes. Dashed horizontal lines correspond to observed covariates residualized-*R*^2^.

Among WW, increased GReX of *MCM10, FAM64A, CCNB2*, and *MMP1* were associated with higher CRS while increased GReX of *VAV3, PCSK6*, and *GNG11* were associated with lower CRS (**Figure 2A**). Among BW, increased GReX of *MMP1* was associated with lower CRS (Proliferation, ROR-P, but not ROR-S) (**Figure 2A**). **Supplementary Figure S4** shows the nominal differences in eQTL architecture across BW and WW for these genes. In particular, for *MMP1*, we found differences in the standardized effects across WW and BW: a sizable proportion of shared eQTLs had discordant effects across WW and BW (**Supplementary Figure S5**). The LD structure for eQTLs differed across WW and BW, with eQTL effect size peaks (-log_10_ p-values: 4.73 (WW); 3.17 (BW)) at differing genomic locations (**Supplementary Figure S5**).

Briefly, to contextualize the functions of these GReX-prioritized genes, *MCM10* is involved in DNA replication, *FAM64A* and *CCNB2* are implicated in progression and regulation of the cell cycle, and *MMP1*, like the broader *MMP* family, is involved in the breakdown of the extracellular matrix (51-55). *GNG11* and *VAV3* are involved in signal transduction: *GNG11* as a component of a transmembrane G-protein and *VAV3* as a guanine nucleotide exchange factor for GTPases (56,57).

Associations between tumor expression of GReX-prioritized genes and CRS were concordant, in terms of direction of association to germline-only effects among WW; findings were discordant among BW where higher tumor expression of MMP1 was associated with higher CRS (**Table 1, Supplementary Table S4**). We found differences in the pattern of associations between genetic ancestry and race with tumor expression and GReX of GReX-prioritized genes (**Supplementary Figure S6**). For instance, while higher African ancestry was associated with higher tumor expression of *MCM10*, higher African ancestry was instead associated with lower GReX of *MCM10*.

**Table 1:** Race-specific associations between germline-regulated tumor gene expression (GReX) of GReX-prioritized genes and CRS. Effect estimates correspond to change in CRS per 1 standard deviation increase in GReX, adjusted for age, estrogen receptor status, stage, and CBCS study phase. 95% confidence intervals of effect sizes are provided. All GReX-prioritized gene and CRS pairs shown here showed overall association FDR-adjusted *P* < 0.10, and FDR-adjusted permutation *P* < 0.05 (across 5,000 permutations of the SNP-gene weights). We also provide signatures that include these genes as reference (**Supplementary Table S1**).

| Gene | Signature | WW (N = 1,043) |  |  | BW (N = 1,083) |  |  |
| --- | --- | --- | --- | --- | --- | --- | --- |
|  |  | ROR-S | Proliferation | ROR-P | ROR-S | Proliferation | ROR-P |
| **MCM10** | IGF | 3.03<br>(1.73, 4.33) | 0.06<br>(0.03, 0.08) | 3.11<br>(1.72, 4.50) | - | - | - |
| **FAM64A** | IGF | 2.57<br>(1.28, 3.86) | 0.05<br>(0.02, 0.07) | 2.64<br>(1.26, 4.02) | - | - | - |
| **CCNB2** | Estradiol | 2.69<br>(1.40, 3.98) | 0.05<br>(0.02, 0.08) | 2.71<br>(1.33, 4.09) | - | - | - |
| **MMP1** | Estradiol | 2.73<br>(1.45, 4.01) | 0.05<br>(0.02, 0.07) | 2.58<br>(1.21, 3.96) | -1.84<br>(-3.12, -0.56) | -0.04<br>(-0.07, -0.02) | -2.21<br>(-3.56, -0.87) |
| **VAV3** | Other | -2.22<br>(-3.51, -0.93) | -0.04<br>(-0.07, -0.02) | -2.40<br>(-3.79, -1.03) | - | - | - |
| **PCSK6** | IGF | -2.16<br>(-3.45, -0.88) | -0.03<br>(-0.06, 0.00) | -1.88<br>(-3.25, -0.50) | - | - | - |
| **GNG11** | Claudin-low | -1.27<br>(-2.56, 0.02) | -0.02<br>(-0.05, 0.00) | -1.42<br>(-2.80, -0.05) | - | - | - |

### Permutation testing provides context to GReX-prioritized gene and CRS associations

To assess the statistical significance for the observed variance in CRS explained by significant GReX-prioritized genes, we conducted two permutation analyses. First, we assessed the per-gene significance of the GReX-CRS associations, conditional on the SNP-trait effects at the locus, by generating a null distribution for the GReX-CRS association via shuffling the SNP-gene weights from the predictive models 5,000 times. We generated a permutation P-value for the GReX-CRS association by comparing to this null distribution. Here, we found that all GReX-CRS associations showed significance in permutation testing at FDR-adjusted P < 0.05 (**Table 1**). These per-GReX-prioritized gene permutation tests show that GReX (of GReX-prioritized genes) adds more context beyond the genetic architecture at the locus and provide evidence that germline genetics to GReX-prioritized gene expression relationship mediates, to some level, the complex genetic effects on CRS.

Next, we quantified the percent variance explained of CRS by the GReX-prioritized genes, in aggregate, by calculating the model adjusted-R^2^ for a regression of covariate-residualized CRS on GReX all GReX-prioritized genes. To context these model adjusted-R^2^, we conducted two permutation tests. First, we permuted the sample labels for covariate-residualized CRS 10,000 times and computed the model adjusted R^2^ at each iteration to generate a null distribution for adjusted R^2^ between GReX-prioritized genes and CRS. Across WW and BW, the observed R^2^ of GReX-prioritized genes against CRS (7-10% among WW and 1% among BW) were statistically significant against the respective null distributions (P-values and distributions in **Figure 2B**).To further contextualize the proportion of variance in CRS explained by GReX-prioritized genes, we computed race-specific heritability estimates using GCTA (58). Given the limited sample size for which CRS data were available, we computed the heritability based on typed SNPs. Moreover, heritability estimates for CRS were stratified by race. Among WW, heritability ranged from 0.13 (SE: 0.23) for ROR-S to 0.21 (SE: 0.23) for Proliferation score. Among BW, heritability was much lower and ranged from 0.01 (SE: 0.12) for Proliferation score to 0.02 (SE: 0.14) for ROR-P. However, we note that heritability estimates from GCTA were imprecise due to limited sample size. Permutation tests for analyses of tumor expression of GReX-prioritized genes and CRS are available in **Supplementary Figure S7**.

Second, we wanted to assess if the GReX of these sets of GReX-prioritized genes (7 in WW and 1 in BW) explained more of the variance in CRS than the GReX of a randomly selected set of genes of the same size. Previous studies have shown that the tumor expression of a set randomly selected genes is likely to be predictive of breast cancer outcomes; we wished to investigate this phenomenon on the GReX level (59,60). Over 10,000 repetitions, we randomly selected 7 and 1 genes in WW and BW subjects, respectively, ran a multivariable regression, and calculated the model adjusted-R^2^ to generate another null distribution. Here again, we found that the true model R^2^ outperformed the null distribution, all showing permutation P < 0.05 in these settings (**Figure 2B**). These permutation tests show that our GReX-prioritized genes, taken together, appreciably explain differences in CRS.

### Associations between GReX-prioritized genes and PAM50 subtype correlations and gene expression

As CRS are constructed from PAM50 subtype-specific correlations and gene expression profiles, we further studied associations between GReX of GReX-prioritized genes and PAM50 SCCs and gene expression to understand how PAM50 subtype and gene expression mediate GReX-prioritized gene and CRS associations. Among WW, a one standard deviation increase in *FAM64A* and *CCNB2* GReX resulted in significantly increased Basal-like SCC while an identical increase in *VAV3, PCSK6*, and *GNG11* GReX resulted in significantly increased Luminal A SCC. The magnitude of increase in correlation for respective subtypes per GReX-prioritized gene was approximately 0.05, and most estimates had credible intervals that did not intersect the null. Among WW, associations between HER2-like SCC and GReX-prioritized genes followed similar patterns to associations for the Basal-like subtype, although associations for HER2 were more precise (**Figure 3A**). We found predominantly null associations between GReX-prioritized genes and Luminal B SCC among WW (**Figure 3A**). Unlike in WW, for BW, an increase in *MMP1* GReX was not associated with Luminal A, HER2 or Basal-like SCCs. Instead, among BW, *MMP1* GReX was significantly negatively associated with Luminal B SCC. Estimates from univariate regressions are provided in **Supplementary Tables S5-S8**.

**Figure 3.**
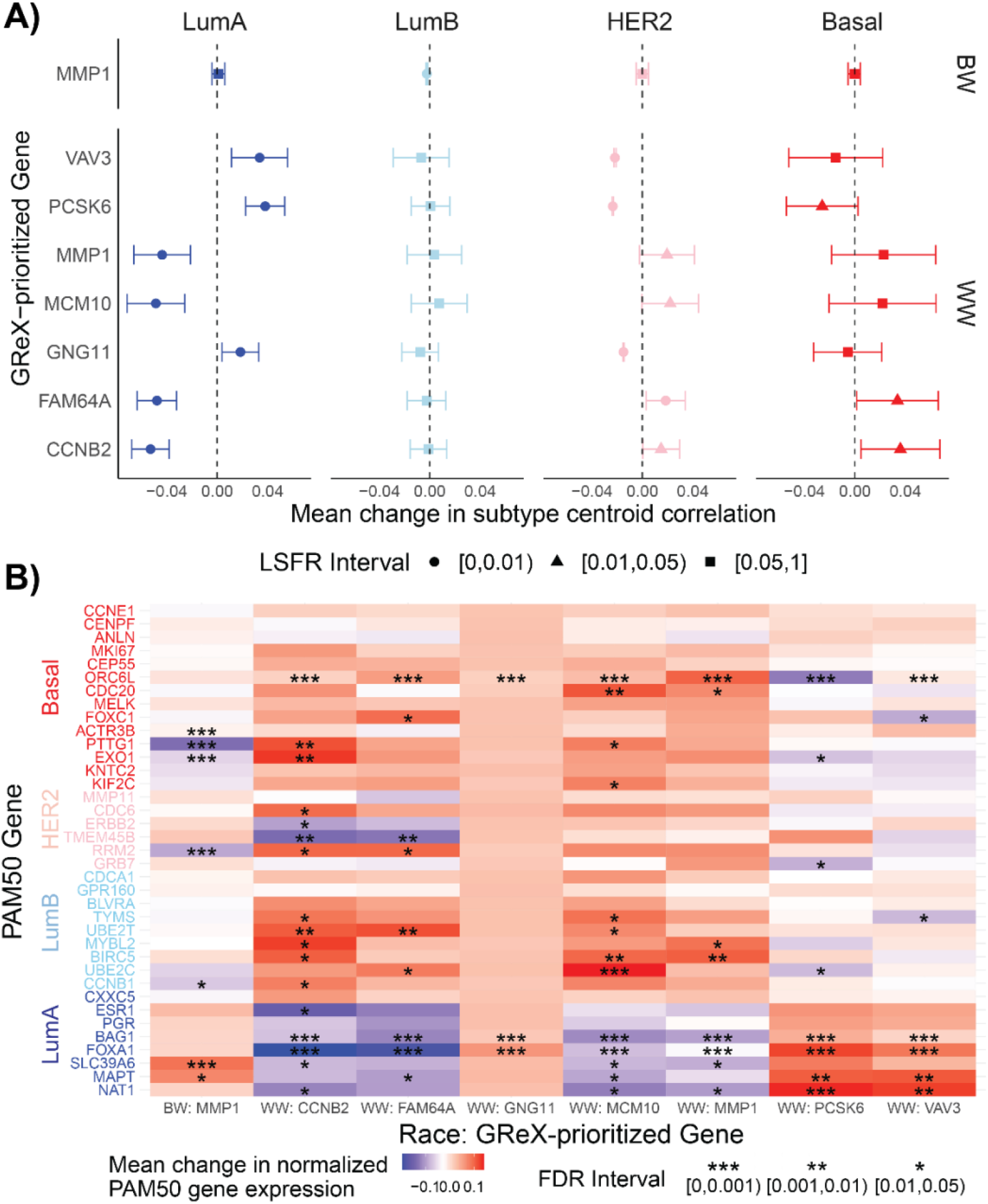
Associations between GReX-prioritized genes and PAM50 SCCs and gene expression. A) Among BW (top) and WW (bottom), associations between GReX-prioritized genes and PAM50 SCCs using Bayesian multivariate regression and multivariate adaptive shrinkage. Effect estimates show change in SCCs (range -1 to 1) for one standard deviation increase in GReX-prioritized gene GReX. Circle, triangle, and square denote corresponding LFSR intervals for effect sizes. B) Heatmap of change in log_2_ normalized PAM50 tumor expression for one standard deviation increase in GReX- prioritized gene GReX. *, **, *** denote FDR intervals for effect sizes.

For both WW and BW, the pattern of associations between GReX-prioritized genes and PAM50 tumor expression were predominantly congruent with observed associations between GReX-prioritized genes and PAM50 SCCs as well as GReX-prioritized genes and CRS (**Figure 3, Supplementary Tables S9-S12**). In WW, a one standard deviation increase in *CCNB2* GReX was associated with significantly increased *ORC6L, PTTG1*, and *KIF2C* (Basal-like genes) expression and *UBE2T* and *MYBL2* (LumB genes) expression. By contrast, a one standard deviation increase in *PCSK6* GReX significantly increased *BAG1, FOXA1, MAPT*, and *NAT1* (LumA genes) expression (**Figure 3B**). While increased *MMP1* GReX was associated with significantly increased expression of *ORC6L* (basal-like gene), *MYBL2*, and *BIRC5* (LumB genes) among WW, this was not the case among BW. Instead, increased *MMP1* GReX among BW was significantly associated with increased expression of *SLC39A6* (LumA gene) and decreased expression of *ACTR3B, PTTG1*, and *EXO1* (Basal-like genes) (**Figure 3B**). Associations between GReX-prioritized genes and PAM50 genes provide a granular, gene interaction level view into the mediation of the GReX-prioritized gene and CRS association, suggesting that *trans*-regulation of subtype-specific PAM50 genes by GReX-prioritized genes in breast tumors could be a possible contributor to subtypes and, subsequently, CRS and recurrence.

## DISCUSSION

Through a GReX analysis, we identified 7 and 1 genes among WW and BW, respectively, for which genetically-regulated breast tumor expression was associated with CRS and underlying PAM50 gene expression and subtype. Among WW, these 7 GReX-prioritized genes explained between 7-10% of the variation in CRS, a large and statistically significant proportion of variance. Among BW, the singular GReX prioritized gene explained a statistically significant ∼1% of the variation in Proliferation score and ROR-P. The magnitudes of these estimates were concordant with race-specific heritability estimates for CRS (13-21% for WW; 1-2% or BW) in this study population and suggest higher germline genetic contribution to CRS among WW compared to BW and as substantial contribution of GReX-prioritized genes to race-specific CRS heritability. There are two key novel aspects to this study. First, existing literature on associations between tumor gene expression and recurrence (for which CRS are a proxy) cannot distinguish between genetic and non-genetic components of effect (61), whereas, here, we estimate the contribution of the genetic component. Second, GReX analysis allows directional interpretation of observed associations that are not possible when correlating tumor gene expression and recurrence. For instance, prior studies report *CCNB2* is upregulated in triple-negative breast cancers (TNBC) but were unable to determine whether increased *CCNB2* expression contributes to development or maintenance of TNBC or is part of the molecular response to cancer progression (62,63). By contrast, GReX is a function of only genetic variation. As such, we can confidently rule out that differences in *CCNB2* GReX are not direct consequences of subtype (and by extension recurrence); however, our observed associations of *CCNB2* GReX and subtype suggest a potential directional relationship for further study. Thus, GReX analysis allows a directional, potentially causal interpretation, subject to effective control for population stratification, minimal horizontal pleiotropy, and assumptions of independent assortment of alleles (22,23).

Our GReX-prioritized gene and subtype associations among WW are consistent with literature on the association between tumor (i.e., genetic and non-genetic) expression of our GReX-prioritized genes and subtype. Prior investigations in cohorts of primarily European ancestry have reported that *MCM10, FAM64A*, and *CCNB2* expression is higher in ER-negative compared to ER-positive tumors (62-64). In studies that compared triple-negative and non-triple negative subtypes, higher *MCM10, FAM64A*, and *CCNB2* expression was detected in triple-negative breast cancer (62,63). Histologically, HER2-enriched and Basal-like subtypes are typically ER-negative, and triple-negatives are similar to Basal-like subtypes (9,65). Moreover, our findings among WW that GReX of *PCSK6* and *VAV3* associated with LumA subtype and LumA-specific gene expression are also consistent with previous results of *PCSK6* and *VAV3* upregulation in ER-positive subtypes (66,67). Importantly, our associations suggest directional mechanisms: from germline variation, to GReX of GReX-prioritized gene, and ultimately, to subtype. Presently, little is known about germline genetic regulation of PAM50 tumor expression. In CBCS, we found that tumor expression of most PAM50 genes is not *cis*-heritable (18). Instead, observed GReX-prioritized gene and PAM50 gene expression associations may implicate *trans*-gene regulation of the PAM50 signature. For instance, we found that *VAV3* GReX is significantly positively associated with tumor expression of *BAG1, FOXA1, MAPT*, and *NAT1* and nominally with increased tumor *ESR1* expression, all of which correspond well with LumA signature. Such *trans*-genic regulation signals, especially in the case of *ESR1*, pose significant clinical and therapeutic implication if confirmed under experimental conditions. For example, *VAV3* has been shown to activate *RAC1*, which upregulates *ESR1* (68,69), but such mechanistic evidence is sparse for other putative GReX-prioritized gene to PAM50 associations. More generally, two of the GReX-prioritized genes among WW have been found to activate transcription factors; *FAM64A* enhances oncogenic nuclear factor-kappa B (NF-κB) signaling while both *FAM64A* and *PCSK6* activate oncogenic *STAT3* signaling (70-72).

Interestingly, we found *MMP1* GReX has divergent associations with CRS across race. There are a few potential explanations. While heritability and proportion of variance in MMP1 expression were similar across WW and BW predictive models, we found that the range of *MMP1* GReX was manifold among WW than BW. Potential differences in influence of germline genetics on tumor expression and CRS by race could be an artifact of divergent somatic or epigenetic factors that CBCS has not assayed (73-76). Second, while studies generally report that *MMP1* tumor expression is higher in triple-negative and Basal-like breast cancer, one study reported that *MMP1* expression in tumor cells does not significantly differ by subtype (77-79). Instead, Bostrom *et al*. reported that *MMP1* expression differs in stromal cells of patients with different subtypes (79). There is evidence to suggest that tumor composition, including stromal and immune components, may influence breast cancer progression in a subtype-specific manner. Future studies should consider expression predictive models that integrate greater detail on tumor cell-type composition to disentangle potential race-specific tumor composition effects on race-specific GReX associations (80,81).

In this study, race (derived from self-report) captures genetic ancestry and additionally, socioeconomic context. Prior investigations using CBCS data have reported concordance between self-reported race and the first principal component of the combined (i.e. WW and BW) genotype matrix. In our analysis of local-ancestry derived global ancestry estimates and self-reported race, we found a similar, high level of concordance. In the absence of available methods that allow stratification or adjustments based on genetic ancestry across the GReX analytic framework, the use of race as a stratifying variable is intended to serve as a proxy for stratification by genetic ancestry. We acknowledge the limitation that race may not be a viable proxy across other populations outside CBCS, and that it is challenging to parse effects seen across race into effects of genetic ancestry and effects of socioeconomic context.

We found marked differences in the pattern of associations between genetic ancestry and race with tumor expression and GReX of GReX-prioritized genes, highlighting potential differences in contributions of germline and non-germline components to tumor expression across European and African ancestry groups. One particular example is *MCM10*. In the literature, higher *MCM10* tumor expression is correlated with Basal-like subtype, which is more prevalent among BW. The spectrum of our observations suggest that higher *MCM10* tumor expression is associated with Basal-like subtype across both BW and WW, but that the germline-regulated component of this expression may be stronger among WW. Similar patterns were seen for *FAM64A* and *CCNB2*. Analyses by race instead of genetic ancestry yielded associations similar in magnitude and direction. Racial differences in non-germline components of tumor expression, including tumor methylation and somatic alternations, may partly explain race-specific differences in GReX-prioritized genes (18,73-76,82,83). Other factors that warrant further investigation include potential greater contribution of *trans*-regulation in tumor gene expression in BW (methods for capturing *trans*-regulation in gene expression predictive models are not as well-developed as those for *cis*-regulation) (18). These factors should be investigated further as transcriptomic and epigenomic datasets for racially-diverse cohorts of breast cancer patients become available.

There are a few limitations to this study. First, as CBCS used a Nanostring nCounter probeset for mRNA expression quantification of genes relevant for breast cancer, we could not analyze the whole human transcriptome. While this probeset may exclude several *cis*-heritable genes, CBCS contains one of the largest breast tumor transcriptomic datasets for Black women, allowing us to build well-powered race-specific predictive models, a pivotal step in ancestry-specific GReX analysis. Second, CBCS lacked data on somatic amplifications and deletions, inclusion of which could enhance the performance of predictive models of tumor expression (84). Third, as recurrence data was collected in a small subset with few recurrence events, we were unable to make a direct comparison between CRS and recurrence results, which may affect clinical generalizability. However, to our knowledge, CBCS is the largest resource of PAM50-based CRS data.

Our analysis provides evidence of race-specific putative germline associations to CRS, mediated through associations between genetically-regulated tumor expression of GReX-prioritized genes and PAM50 expressions and subtype. This work underscores the need for larger and more diverse cohorts for genetic epidemiology studies of breast cancer. Future studies should consider subtype-specific genetics (i.e., stratification by subtype in predictive model training and association analyses) to elucidate heritable gene expression effects on breast cancer outcomes both across and within subtype, which may yield further hypotheses for more fine-tuned clinical intervention.

## Data Availability

Expression data from CBCS is available on NCBI GEO with accession number GSE148426. CBCS genotype datasets analyzed in this study are not publicly available as many CBCS patients are still being followed and accordingly CBCS data is considered sensitive; the data is available from M.A.T upon reasonable request. Supplementary Data includes summary statistics for eQTL results, tumor expression models, and relevant R code for training expression models in CBCS and are freely available at https://github.com/bhattacharya-a-bt/CBCS_TWAS_Paper/. R code for analyses provided in this paper are available at https://github.com/APUNC/CBCS---Risk-of-Recurrence-Paper.

http://bcac.ccge.medschl.cam.ac.uk/bcacdata/icogs-complete-summary-results

https://github.com/APUNC/CBCS---Risk-of-Recurrence-Paper

https://github.com/bhattacharya-a-bt/CBCS_TWAS_Paper/

http://bcac.ccge.medschl.cam.ac.uk/bcacdata/icogs-complete-summary-results

https://github.com/APUNC/CBCS---Risk-of-Recurrence-Paper

## ABBREVIATIONS

BW: Black Women
CBCS: Carolina Breast Cancer Study
CRS: Continuous Risk of recurrence Score
eQTL: expression Quantitative Trait Locus
ER: Estrogen Receptor
FDR: False Discovery Rate
GReX: Genetically-Regulated tumor eXpression
GWAS: Genome-Wide Association Study
HR: Hormone Receptor
LFSR: Local False Sign Rate
LumA: Luminal A
LumB: Luminal B
NC: North Carolina
ROR: Risk of Recurrence
SCC: Subtype-Centroid Correlations
SNP: Single Nucleotide Polymorphism
TCGA: The Cancer Genome Atlas
TWAS: Transcriptome-Wide Association Study
WW: White Women

## ACKNOWLEDGEMENTS

We thank the Carolina Breast Cancer Study participants and volunteers. We also thank Colin Begg, Jianwen Cai, Katherine Hoadley, Yun Li, and Bogdan Pasaniuc for valuable discussion during the research process. We thank Erin Kirk and Jessica Tse for their invaluable support during the research process. We thank the DCEG Cancer Genomics Research Laboratory and acknowledge the support from Stephen Chanock, Rose Yang, Meredith Yeager, Belynda Hicks, and Bin Zhu.

## AUTHOR CONTRIBUTIONS

Conceptualization: AP, MAT, MIL, AB. Data curation: MG, AFO, CMP, MAT. Formal analysis: AP, MAT, MIL, AB. Funding acquisition: AP, MG, AFO, CMP, MAT, MIL. Methodology: AP, MIL, AB. Project administration: MAT, MIL, AB. Resources: MG, AFO, CMP, MAT, MIL. Supervision: MAT, MIL, AB. Visualization: AP, AB. Writing – original draft: AP, AB. Writing – reviewing and editing: AP, MG, AFO, CMP, MAT, MIL, AB.

## AVAILABILITY OF DATA AND MATERIALS

Expression data from CBCS is available on NCBI GEO with accession number GSE148426. CBCS genotype datasets analyzed in this study are not publicly available as many CBCS patients are still being followed and accordingly CBCS data is considered sensitive; the data is available from M.A.T upon reasonable request. Supplementary Data includes summary statistics for eQTL results, tumor expression models, and relevant R code for training expression models in CBCS and are freely available at https://github.com/bhattacharya-a-bt/CBCS_TWAS_Paper/. Scripts utilized in this analysis are provided at https://github.com/APUNC/CBCS---Risk-of-Recurrence-Paper.

