## Supplementary Materials for "Gene-level germline contributions to clinical risk of recurrence scores in Black and White breast cancer patients"

This document contains Supplementary Methods, References, and Figures S1-S6. Supplementary Tables S1-S12 are provided in a separate .xlsx file due to their size.

#### 1 Supplementary Methods

Here, we provide mathematical details of computational methods used in this analysis.

##### 1.1 eQTL analysis

We assessed the additive relationship between the log-transformed gene expression values for a gene $g$ and genotypes with linear regression analysis using MatrixeQTL [1]:

$$E_g = X_s\beta_s + X_C\beta_C + \epsilon_g,$$

where $E_g$ is the log-transformed gene expression values for given gene $g$, $X_s$ is the vector of genotype dosages for a SNP $s$, $C$ is a matrix of covariates, $\beta_s$ and $\beta_C$ are the effect sizes on gene expression for the SNP $s$ and covariates $C$, respectively. $\epsilon$ is assumed to be random error such that $\epsilon \sim N(0, \sigma^2)$, with common variance $\sigma^2$ across all genes.

We calculate both *cis*- (variant-gene distance less than 500 kilobases) and *trans*-eQTLs between all variants and all genes. We tested for the significant of $\beta_s$ with a two-sided Wald test of the null hypothesis $H_0 : \beta_S = 0$. We conducted all eQTL analyses stratified by race and controlling for the following covariates: age, BMI, postmenopausal status, and the first 5 principal components of the joint Black (AA) and White (WW) genotype matrix. We exclude samples with Normal-like subtype, as classified by the PAM50 classifier [2], due to generally low tumor content.

##### 1.2 Tumor gene expression imputation models

Genes with significantly *cis*-heritable tumor expression were prioritized for predictive model training, adopted from PrediXcan and FUSION [3, 4]. We estimated eQTL-effect sizes for tumor expression from germline variant in the following process.

First, gene expression was residualized for the covariates $C$ included in the eQTL models (age,

BMI, postmenopausal, and genotype PCs for population stratification) given the following ordinary least squares (OLS) model:

$$E_g = X_C \beta_C + \epsilon_g.$$

We define the covariate-residualized $\tilde{E}_g \equiv E_g - X_C \hat{\beta}_C$, where $\hat{\beta}_C$ is the OLS estimator.

For a given gene $g$, we consider the following linear predictive model, stratified by race:

$$\tilde{E}_g = X_g w_g + \epsilon_g,$$

where $X_g$ is the genotype matrix for gene $g$ that includes all *cis*-SNPs within 500 kb of either the 5' or 3' end of the gene, $w_g$ is a vector of effect sizes for eQTLs in $X_g$, and $\epsilon_g$ is Gaussian random error. We estimate $w_g$ with the best predictive of three schemes:

1. elastic-net regularized regression with mixing parameter $\alpha = 0.5$ and $\lambda$ penalty parameter tuned over five-fold cross-validation [5];
2. linear mixed modeling where the genotype matrix $X_g$ is treated as a matrix of random effects and $\hat{w}_g$ is taken as the best linear unbiased predictor (BLUP) of $w_g$ using rrBLUP [6]; or
3. multivariate linear mixed modeling using GEMMA [7].

In these models, the genotype matrix $X_g$ is pruned for linkage disequilibrium prior to modeling using a window size of 50, step size of 5, and LD threshold of 0.5 using PLINK [8] to account for redundancy in signal. These LD-pruning thresholds and window sizes are not stringent [9] and leads to greater five-fold cross-validation $R^2$ [10].

To impute into external cohorts, we construct the germline genetically-regulated tumor expression $\text{GReX}_g$ given $\hat{w}_g$ in the predictive model:

$$\text{GReX}_g = X_{g,\text{new}} \hat{w}_g,$$

where $X_{g,\text{new}}$ is the genotype matrix of all available SNPs in the feature set of $\hat{w}_g$ in a GWAS cohort. As we impute GReX in CBCS, for individuals that were used in the training data set, we impute their GReX via cross-validation to avoid data leakage. We use $\text{GReX}_g$ (scaled to zero mean and unit variance 1) as a primary predictor of interest in multiple linear models for various outcomes: risk of recurrence or proliferation scores, recurrence time-to-event outcomes, etc.

#### 1.3 Bayesian analysis of correlated phenotypes

We detected several genes with GReX associated with at least one of ROR-P, ROR-S, and proliferation scores, which are functions of gene expressions that determine PAM50 molecular subtypes [2]. We wished to detect whether any of these GReX are associated with the tumor expression of PAM50 genes. In essence, here, we are conducting a trans-eQTL mapping using strategies similar to Wheeler *et al*'s trans-PrediXcan [11] and Liu *et al*'s GBAT methods [12]. However, instead of an univariate approach that cannot take into account the dense correlation structure between the expression of these genes, we consider a multivariate linear model with correlated outcomes:

$$\mathbf{Y} = \mathbf{XB} + \mathbf{E},$$

where $\mathbf{Y}$ is an $n \times m$ matrix for $n$ samples (on rows) and $m$ outcomes (columns), $\mathbf{X}$ is an $n \times k$ matrix of $k$ predictors (columns), and $\mathbf{B}$ is a $k \times m$ matrix of effect sizes for each of the $k$ predictors. The matrix $\mathbf{E}$ represents the correlated errors with a multivariate normal distribution, such that the vector of errors for a given observation is correlated: $\epsilon_i \sim N(0, \mathbf{\Sigma}_\epsilon)$. This imposes correlation between outcomes in the regression model. We approach estimation of $B$ and $\mathbf{\Sigma}_\epsilon$ through a straight-forward Bayesian multivariate regression with conjugate priors [13]. We assume the following prior distributions:

$$\begin{aligned} \mathbf{\Sigma}_\epsilon &\sim \mathcal{W}^{-1}(\mathbf{V}_0, \nu_0) \\ \beta|\mathbf{\Sigma}_\epsilon &\sim N\left(\beta_0, \mathbf{\Sigma}_\epsilon \otimes \mathbf{\Lambda}_0^{-1}\right), \end{aligned}$$

where $\mathcal{W}^{-1}(\mathbf{A}, b)$ represents an inverse Wishart distribution with scale matrix $\mathbf{A}$ and $b$ degrees of freedom and $\otimes$ represents the Kronecker product operator. Now, the posterior joint distribution of $\beta$ and $\mathbf{\Sigma}_\epsilon$ is:

$$\begin{aligned} p\left(\beta, \mathbf{\Sigma}_\epsilon|\mathbf{Y}, \mathbf{X}\right) \propto |\mathbf{\Sigma}_\epsilon|^{-(\nu_0+m+1)/2} &\exp\left\{-\frac{1}{2}\text{tr}\left(\mathbf{V}_0\mathbf{\Sigma}_\epsilon\right)^{-1}\right\} \\ \times |\mathbf{\Sigma}_\epsilon|^{-k/2} &\exp\left\{-\frac{1}{2}\text{tr}\left((\mathbf{B}-\mathbf{B}_0)'\Lambda_0(\mathbf{B}-\mathbf{B}_0)\right)\right\} \\ \times |\mathbf{\Sigma}_\epsilon|^{-n/2} &\exp\left\{-\frac{1}{2}\text{tr}\left((\mathbf{Y}-\mathbf{XB})'(\mathbf{Y}-\mathbf{XB})\mathbf{\Sigma}_\epsilon^{-1}\right)\right\}, \end{aligned}$$

with $\beta_0 = vec(\mathbf{B}_0)$. If we group quadratic forms together with $\mathbf{B}_n = (\mathbf{X}'\mathbf{X} + \mathbf{\Lambda}_0)^{-1}(\mathbf{X}'\mathbf{Y} + \mathbf{\Lambda}_0\mathbf{B}_0)$, we can find that the posterior takes the form of a product of an inverse Wishart distribution and a Matrix normal distribution:

$$\begin{aligned} \mathbf{\Sigma}_\epsilon|\mathbf{Y}, \mathbf{X} &\sim \mathcal{W}^{-1}(\mathbf{V}_n, \nu_n) \\ \mathbf{B}|\mathbf{\Sigma}_\epsilon, \mathbf{Y}, \mathbf{X} &\sim \mathcal{MN}_{k,m}\left(\mathbf{B}_n, \mathbf{\Lambda}_n^{-1}, \mathbf{\Sigma}_\epsilon\right), \end{aligned}$$

where

$$\begin{aligned} \mathbf{V}_n &= \mathbf{V}_0 + (\mathbf{Y} - \mathbf{XB}_n)'(\mathbf{Y} - \mathbf{XB}_n) + (\mathbf{B}_n - \mathbf{B}_0)'\mathbf{\Lambda}_0(\mathbf{B}_n - \mathbf{B}_0) \\ \nu_n &= \nu_0 + n \\ \mathbf{B}_n &= (\mathbf{X}'\mathbf{X} + \mathbf{\Lambda}_0)^{-1}(\mathbf{X}'\mathbf{Y} + \mathbf{\Lambda}_0\mathbf{B}_0) \\ \mathbf{\Lambda}_n &= \mathbf{X}'\mathbf{X} + \mathbf{\Lambda}_0. \end{aligned}$$

We call this extension *Bayesian trans-QTL mapping*, or BtQTL for short.

##### 1.3.1 Simulation study

We compared BtQTL against GBAT [12] with correlated gene expression through simulations.

Here, we based simulated data off expression and genotype data from CBCS. We randomly selected a gene (call it $g_c$) from Chromosome 22, extracted the *cis*-SNPs within a 500 kb window, and used the linkage disequilibrium structure from these SNPs to simulate $n$ genotypes. We then simulated eQTL effect sizes with a causal proportion $p_c$ of *cis*-eQTLs and simulated an expression vector for the *cis*-gene expression with scaled effect sizes such that the variance explained from *cis*-genotypes is $h_c^2$ [14]. Next, considering the genes not on Chromosome 22, we selected $t$ genes with a sparse correlation matrix (all absolute cross-gene correlations less than 0.1) or dense correlation matrix (all absolute cross-gene correlations greater than 0.4). We generate an effect size $\beta_t \sim N(0,1)$ for one of the $t$ randomly selected *trans*-genes (call it $g_t$) to simulate a trans-association between $g_c$ and $g_t$ and scale the effect so that $g_c$ explains $h_t^2$ of the variance in expression of $g_t$.

Now, consider the following model:

$$\mathbf{Y} = X\beta + \mathbf{E},$$

where $Y$ is an $n \times t$ matrix of correlated gene expressions, $X$ is the vector of expression for $g_c$, $\beta$ is the vector of effect sizes of expression of $g_c$ on the $t$ trans-genes, and $\mathbf{E}$ represents the random error. Now, to ensure the correlation between columns of $\mathbf{Y}$ reflects the correlation matrix from the observed CBCS data, we match moments to generate a multivariate normal random matrix for $\mathbf{E}$:

Let $\mathbf{C}$ be the observed correlation matrix. Let $Y_i$ and $Y_j$ be the vectors of expression of the $i$th and $j$th genes. We find

$$\begin{aligned} Cov(Y_i, Y_j) &= Cov(X\beta_i + \epsilon_i, X\beta_j + \epsilon_j) \\ &= \beta_i\beta_j Var(X) + Cov(\epsilon_i, \epsilon_j). \end{aligned}$$

Now, for $i \neq j$ and $c_{ij}$ the correlation between $Y_i$ and $Y_j$, we have

$$Cov(Y_i, Y_j) = c_{ij}\sigma_i\sigma_j = \beta_i\beta_j Var(X) + Cov(\epsilon_i, \epsilon_j).$$

We can solve for $Cov(\epsilon_i, \epsilon_j)$, as other values are known, and use these values across $i, j \in \{1, \ldots, t\}$ to simulate $\mathbf{E}$. The $i$th diagonal entry for $\mathbf{E}$ (the variance of $\epsilon_i$) can be taken from the equivalence $\sigma_i^2 = \beta_1^2 Var(X) + Var(\epsilon_i)$. This gives us a simulated $\mathbf{Y}$ with a given correlation matrix to run GBAT and the Bayesian regression approach. We conduct 10,000 simulations across a variety of parameters: $n \in \{200, 500, 1000\}$, $p_c \in \{0.05, 0.10, 0.25\}$, $h_c^2 = 0.15$, $h_t^2 \in \{0.05, 0.10\}$, $t \in \{2, 5\}$, and a sparse (correlations all under 0.2) or dense (correlations all over 0.5) correlation matrix. We apply both the GBAT method and the Bayesian regression model outlined above. For GBAT, we compute a false positive rate for the elements of $\beta$ that are set to 0 and power for nonzero $\beta$ at a Bonferroni corrected significance threshold of $P < 0.05/t$ for $\beta$. For the Bayesian approach, we generate a $(100 - 5/2t)$% credible interval for $\beta$ to gauge false positive rates and power by finding whether the interval includes the null (effect size of 0). **Supplementary Figure S1** shows a comparison of GBAT and BtQTL across these simulation parameters, showing the slight advantage of BtQTL over the GBAT mapping framework, which generally increases with sample size and the number of total *trans*-genes considered. We further highlight the advantage of BtQTL over GBAT with increased number of *trans*-genes considered with the simulation parameters: $n = 500, p_c = 0.10, h_c^2 = 0.15, h_t^2 = 0.10,$ and $t = 30$ (**Supplementary Figure S7**). We vary the number of true *trans*-QTLs from 1, 2, 5, and 10. As the number of the considered genes increases, BtQTL has a larger advantage over the univariate methods (**Supplementary Figure S7**).

[9] Nilanjan Chatterjee, Jianxin Shi, and Montserrat García-Closas. Developing and evaluating polygenic risk prediction models for stratified disease prevention, 7 2016.

[10] Arjun Bhattacharya, Montserrat García-Closas, Andrew F. Olshan, Charles M. Perou, Melissa A. Troester, and Michael I. Love. A framework for transcriptome-wide association studies in breast cancer in diverse study populations. *Genome Biology*, 21(1):42, 12 2020.

[11] Heather E. Wheeler, Sally Ploch, Alvaro N. Barbeira, Rodrigo Bonazzola, Angela Andaleon, Alireza Fotuhi Siahpirani, Ashis Saha, Alexis Battle, Sushmita Roy, and Hae Kyung Im. Imputed gene associations identify replicable trans-acting genes enriched in transcription pathways and complex traits. *Genetic Epidemiology*, 43(6):gepi.22205, 4 2019.

[12] Xuanyao Liu, Joel A Mefford, Andrew Dahl, Yuan He, Meena Subramaniam, Alexis Battle, Alkes L Price, and Noah Zaitlen. GBAT: a gene-based association test for robust detection of trans-gene regulation. *Genome Biology*, 21(1):211, 12 2020.

[13] George C Tiao and Arnold Zellner. On the Bayesian Estimation of Multivariate Regression. Technical Report 2, 1964.

[14] Nicholas Mancuso, Malika K. Freund, Ruth Johnson, Huwenbo Shi, Gleb Kichaev, Alexander Gusev, and Bogdan Pasaniuc. Probabilistic fine-mapping of transcriptome-wide association studies. *Nature Genetics*, 51(4):675–682, 4 2019.

### 3 Supplementary Figures

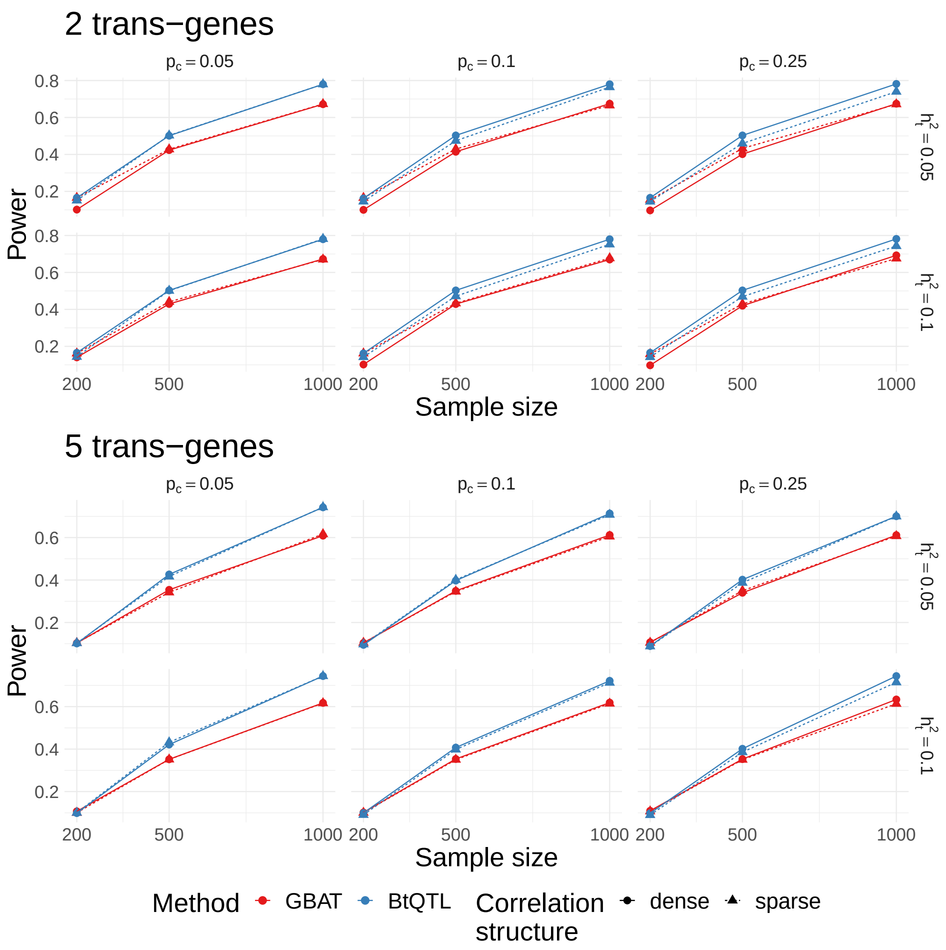


Figure S1: *Power comparison between univariate GBAT and Bayesian trans-QTL mapping with many genes.* We compare the power ($Y$-axis) to detect trans-genetic associations between genetically regulated expression of a gene and 2 (top) and 5 (bottom) genes on different chromosomes using GBAT [12] (red) and BtQTL (blue). The $X$-axis shows the sample size, the causal proportion of *cis*-eQTLs is shown on the horizontal strip labels, and the total distal heritability of the genes is shown on the vertical strip labels. We show differences in power across dense (circles, solid line; absolute correlation between all genes $\geq 0.40$) and sparse (triangle, dotted line; absolute correlation between all genes $\leq 0.10$) correlation structures. Here, we assume only 1 of the 2 or 5 *trans*-genes considered have a truly non-zero association.

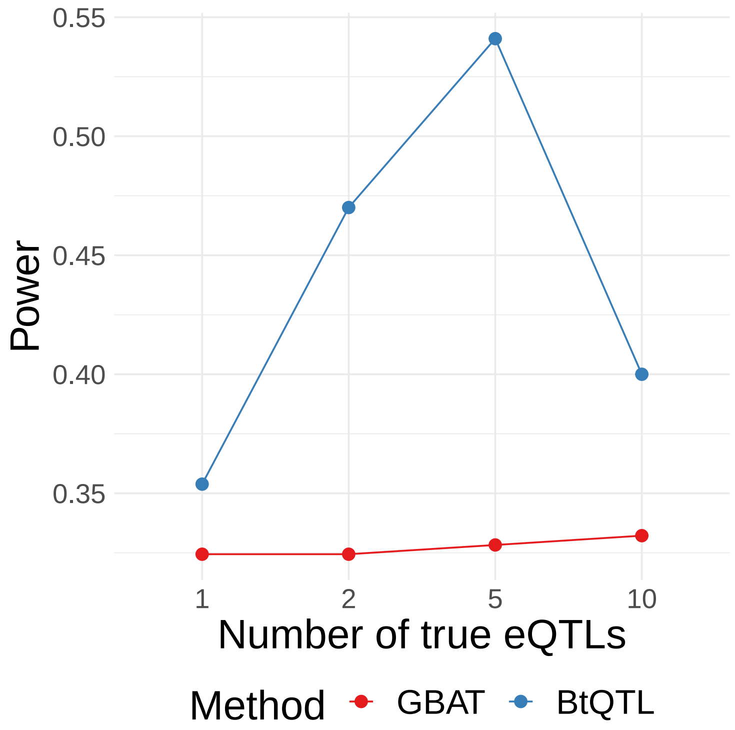


Figure S2: *Power comparison between univariate GBAT and Bayesian trans-QTL mapping.* We compare the power ($Y$-axis) to detect trans-genetic associations between genetically regulated expression of a gene and 30 genes on different chromosomes using GBAT [12] (red) and BtQTL (blue). The $X$-axis shows the number of simulated non-zero *trans*-associations. We assume a sample size of 500, causal proportion of *cis*-eQTLs 0.10, distal heritability of 0.10, and a dense correlation structure.

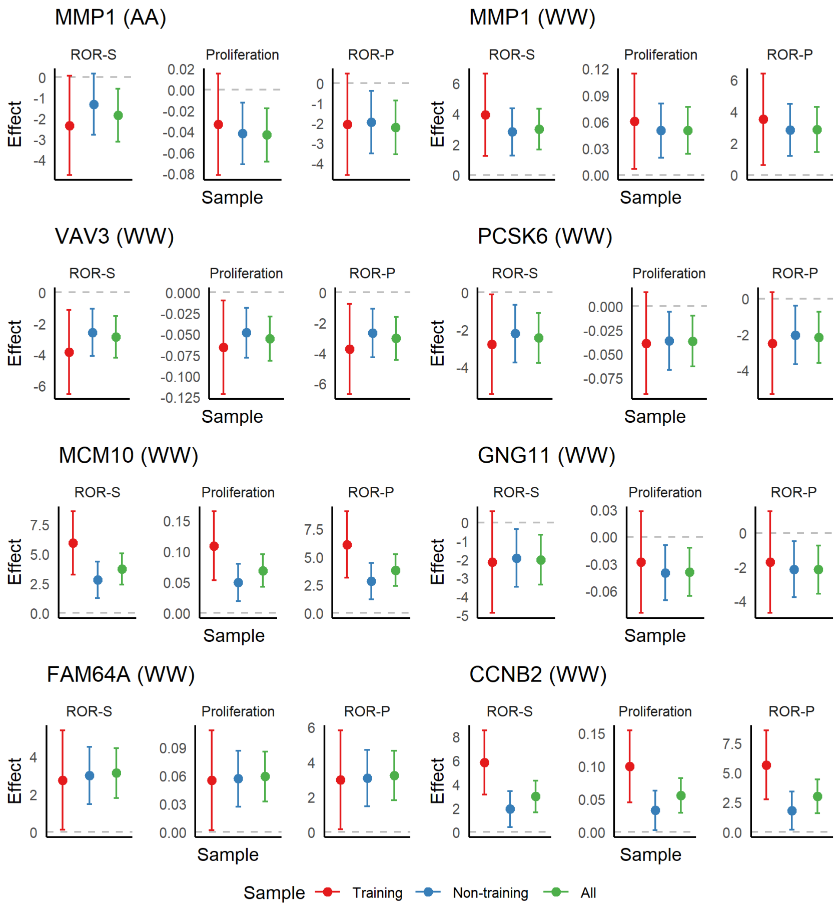


Figure S3: *Sensitivity analysis of GReX-CRS association across training, non-training, and all samples.* We compare the effect size and 95% confidence intervals ($Y$-axis) of the GReX-CRS association across only training samples (blue), only samples not used in training (red), and all samples (green), shown on the $X$-axis. The grey line represents the null association with effect size 0.

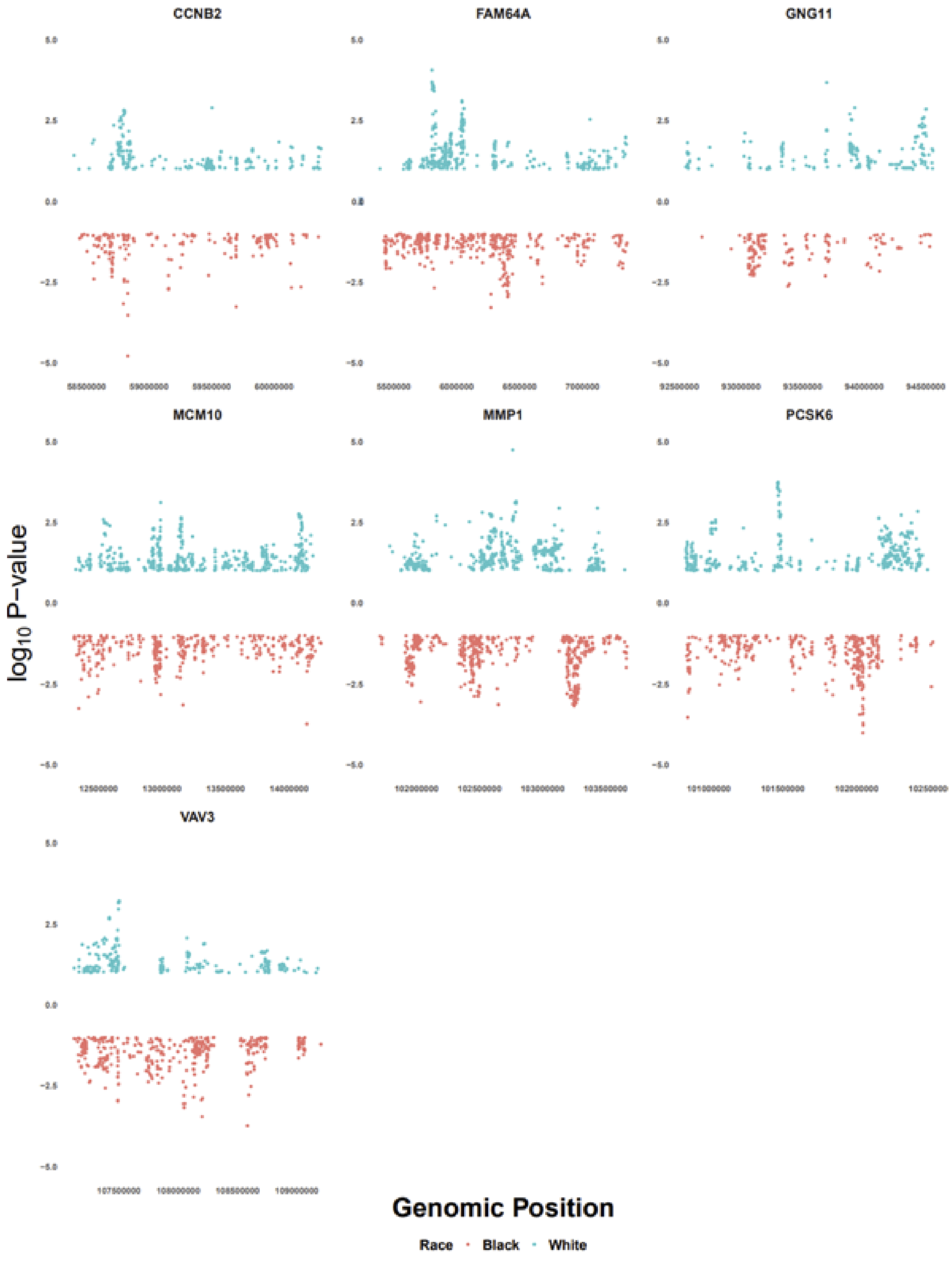


Figure S4: *Effect of cis-genetic variants on tumor expression of GReX-prioritized genes among White women and Black women.* Manhattan plot of cis-eQTLs across Black (red) and White (blue) women.

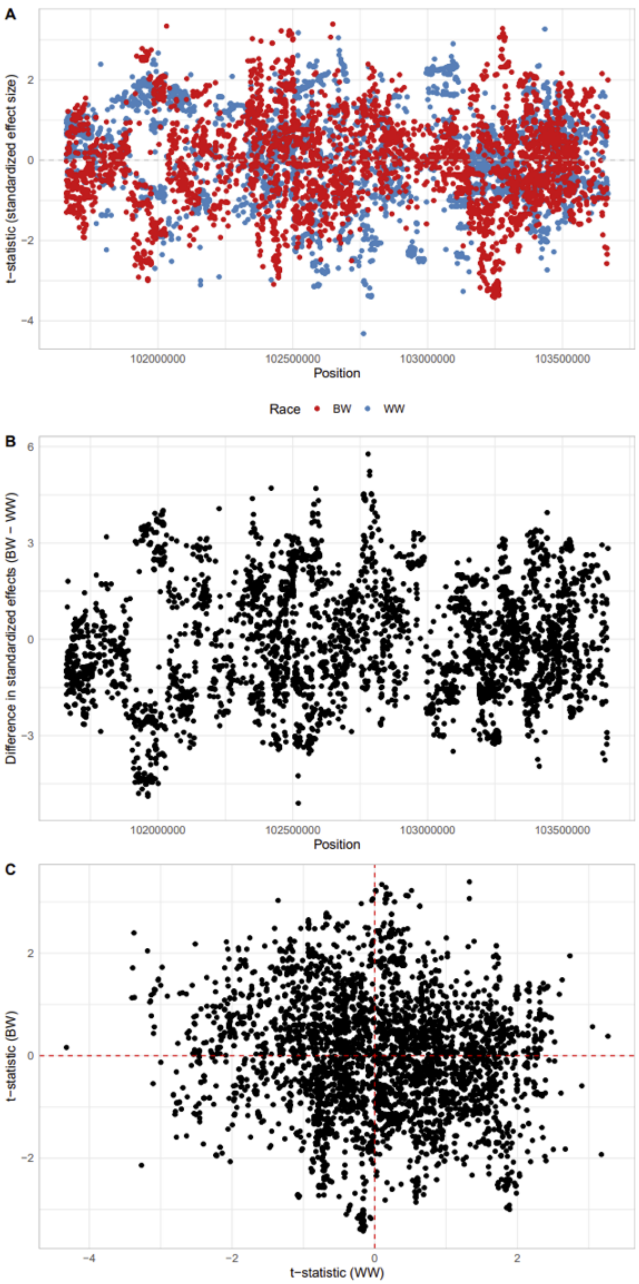


Figure S5: *Comparison of MMP1 eQTLs across Black and White women in CBCS*. (A) Miami plot of standardized effect sizes of eQTLs across BW (red) and WW (blue). (B) Miami plot of difference in standardized effect sizes (BW - WW) of eQTLs across BW and WW women. (C) Scatter plot of standardized effect sizes of eQTLs in WW (X-axis) and BW (Y-axis).

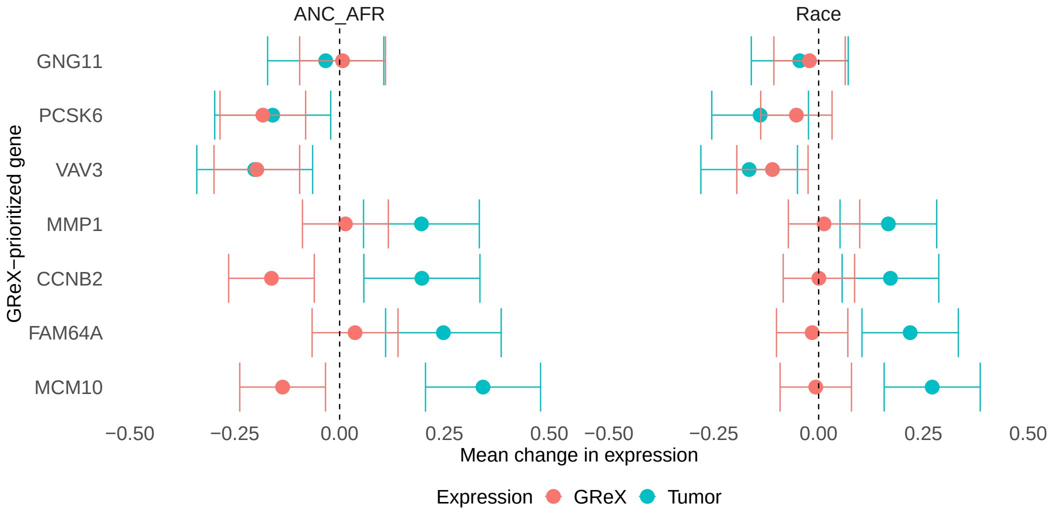


Figure S6: *Associations between tumor expression and GReX of GReX-prioritized genes and genetic ancestry and self-reported race*. Caterpillar plots of effect sizes and standard errors for effect of African ancestry (left) and race (right) on GReX (red) and full tumor expression (blue) for GReX-prioritized genes.

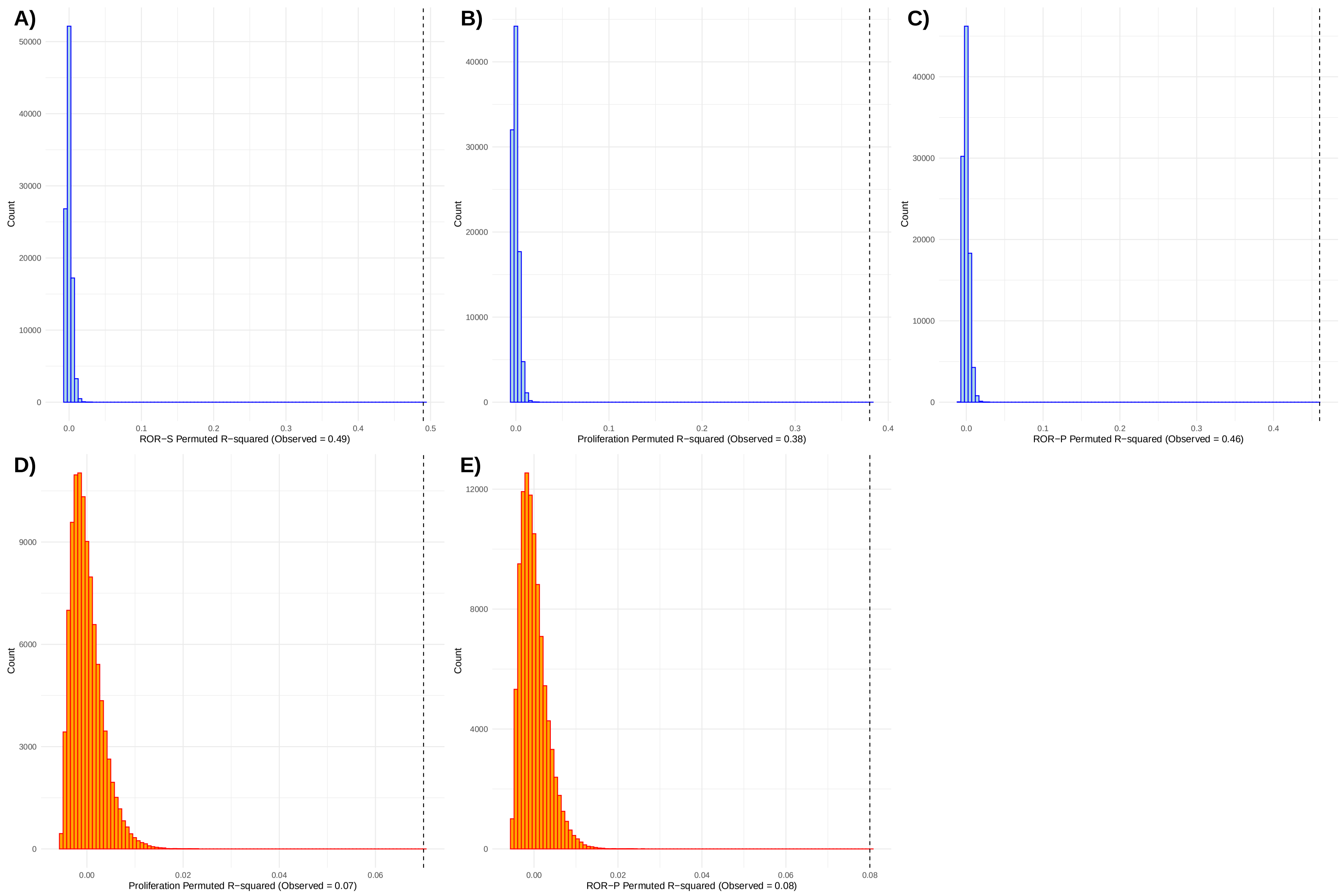


Figure S7: *Histograms of model $R^2$ for GReX models.* Histograms correspond to null distributions of covariates (age at selection, estrogen receptor status, study phase, tumor stage) residualized-$R^2$ for regressions of CRS against BT expression of TWAS-genes. Dashed vertical lines correspond to observed covariates residualized-$R^2$. Light blue denotes WW and orange denotes BW.
